# EEG-fMRI reveals biological motion network disruptions as early markers of Multiple Sclerosis progression

**DOI:** 10.1101/2025.08.25.25334369

**Authors:** Júlia F. Soares, Maria Caranova, Irina Santos, Sónia Batista, Miguel Castelo-Branco, João V. Duarte

## Abstract

Multiple sclerosis (MS) is a demyelinating disease that disrupts neuronal connectivity and alters functional networks. While resting-state approaches are common, task-based paradigms—especially those probing connectivity—remain underexplored. The biological motion (BM) task stands out by engaging highly myelinated, clinically relevant regions and targeting social cognition, a domain often overlooked in MS. Coupling this task with EEG-fMRI increases sensitivity to subtle disease-related changes, making it valuable for tracking progression and treatment effects.

We acquired longitudinal simultaneous EEG-fMRI data during a BM task from 18 early relapsing-remitting MS patients and 18 matched healthy controls at baseline and 10-month follow-up. EEG source activity was constrained using concurrent fMRI priors, and effective connectivity (EC) within the BM network was computed using Dynamic Causal Modelling and Parametric Empirical Bayes. We identified alterations driven by disease (group differences), time (longitudinal changes), and their interaction, and examined their associations with disability, fatigue, and cognitive performance—including social cognition.

We found early disruption of BM network, associated with greater disability and cognitive deficits, particularly in social cognition. While connectivity, disability, and cognition may improve over time—likely due to treatment—new altered connections emerged at follow-up, negatively correlating with cognition. Decreased EC was generally associated with worse function, whereas increased EC—particularly in emotion-related regions—may reflect compensatory mechanisms, though it was also linked to greater fatigue.

The confirmed sensitivity of the combined methods to detect early and subtle alterations in MS underscores their potential to predict cognitive decline and to inform interventions aimed at preserving brain function.

## 1. Introduction

Multiple sclerosis (MS) is a chronic inflammatory disease of the central nervous system and the leading cause of non-traumatic neurological disability in young adults. ^1–5^ It is primarily characterized by demyelination, which disrupts axonal conduction, ^6–8^ and leads to both focal white matter lesions and diffuse damage in normal-appearing tissue, compromising neural connectivity. ^9^ These alterations—along with cortical demyelination, neuroaxonal loss, and grey matter atrophy—contribute to the broad spectrum of motor, cognitive, and sensory impairments in MS, as well as to fatigue, depression, and anxiety. ^2,10,11^. However, these symptoms often lack correspondence with structural MRI findings, supporting the notion that functional neuroimaging biomarkers may provide more sensitive and clinically relevant insights into disease impact, progression, and treatment efficacy. ^12–14^

In this context, functional MRI (fMRI) and EEG, have been widely used to study brain function in MS. ^15–18^ While fMRI offers high spatial resolution, EEG captures fast neural dynamics. Despite its significantly lower cost compared to fMRI, EEG lacks the ability to accurately localize FC changes but given the complementary aspects of EEG and fMRI, their integration has been sought. ^19,20^ When merged, these techniques yield potential to offer a more complete view of brain function, enhancing sensitivity to subtle alterations and improving the assessment of network dysfunction in MS.

Importantly, most studies in MS are conducted in resting-state conditions, ^21^ which may fail to capture task-specific brain network alterations. In contrast, task-based paradigms actively engage cognitive and motor pathways, potentially revealing subtle impairments—particularly in early stages—that resting-state approaches might overlook. ^22,23^ Notably, brain networks display distinct functional properties during task performance compared to rest,^24,25^ supporting the idea that task-based approaches may provide additional insights into clinically relevant dysfunction and the mechanisms through which MS affects brain function.

Furthermore, most of these studies using either fMRI or EEG have focused on brain activation or simple FC measures like correlations, between brain regions which are not specific enough, and may overlook subtle yet meaningful alterations ^21,22,26,27^ In contrast, estimating effective connectivity (EC), allows to explore causal interactions between brain regions, offering deeper insights into how information is processed within neural networks ^28^.One powerful approach for assessing EC is dynamic causal modelling (DCM), a Bayesian statistical method that enables the unveiling of neuronal network activity that originates the observed signals (hemodynamic signals in fMRI data or electromagnetic time-series signals in EEG data). ^29,30^. However, so far, only two studies have use DCM in MS.^31,32^

Importantly, DCM requires a plausible and a priori-defined model of neural interactions to infer how different brain regions influence each other—both in direction and strength—and how these connections are modulated by experimental conditions. In the context of MS, the biological motion (BM) perception paradigm emerges as a potentially informative approach, given that i) is well-established and ecologically valid task, previously validated using DCM by Sokolov et al. ^33^ , and ii) this paradigm consistently engages a distributed network of highly myelinated brain regions,^34,35^ that are involved in social cognition,^33^ iii) it has been recently demonstrated by Bonventre et al.,^36^ in a purely behavioural study without neuroimaging data, that the perception of BM is impaired in individuals in MS when using this same paradigm. Therefore, such approach targeting a highly myelinated network underlying social cognition— given that social cognitive deficits are increasingly recognized in MS yet often overlooked in neuroimaging research,^10,37,38^ —combined with the fact that the perception of BM, using this specific paradigm, has already been shown to be impaired in MS, may increase the likelihood of detecting disease-related functional alterations that conventional approaches might fail to capture, particularly at early stages.

Lastly, most previous studies in MS are cross-sectional, limiting understanding of the temporal dynamics of disease progression. Task-based and resting-state fMRI studies in early MS often report increased activation and connectivity, respectively—commonly interpreted as compensatory mechanisms^22^. In contrast, most EEG studies have focused on event-related potentials (ERPs), typically revealing delayed latencies and reduced amplitudes, indicative of slowed neural processing and reduced functional efficiency.^26,39^ Nonetheless, increased ERP amplitudes have also been observed, potentially reflecting compensatory recruitment.^40,41^ Despite general trends reported in the literature, inconsistencies persist—some studies report increased activation or connectivity, while others report decreases. Moreover, the interpretation of these differences varies widely, with some suggesting adaptive mechanisms and others proposing maladaptive processes.^42–44^ These discrepancies are highly dependent on the specific networks and brain regions investigated, as both the direction of the observed changes and their functional relevance may vary accordingly.^45^ Notably, there is a lack of task-based paradigms specifically designed to probe connectivity, limiting the development of strong formal hypotheses concerning the strength, direction and precise locations of between-groups alterations.

In this study, we address these gaps by presenting, for the first time, a longitudinal analysis of EC using simultaneous EEG-fMRI, through a fMRI-informed reconstruction of EEG sources during performance of a BM task in recently diagnosed patients with early relapsing-remitting MS (RRMS) and matched healthy controls (HC). We aimed to identify potential indicators of disease progression by assessing both cross-sectional and longitudinal changes in EC, using a biologically grounded DCM model based on the well-characterised BM network validated by Sokolov et al., and analysed within the Parametric Empirical Bayes (PEB) framework. Additionally, we explored associations between connectivity strength, physical disability, and cognitive performance to better understand the functional relevance of EC alterations.

Although primarily exploratory, we hypothesised 1) that this multimodal approach— combining simultaneous EEG-fMRI, a robust connectivity model, and a task engaging a highly myelinated and clinically relevant network—would allow for the detection of very early and subtle functional alterations in MS and their evolution over time, which would correlate with clinical and cognitive symptoms, particularly those related to social cognition. The potential for early detection is further supported by the inclusion of recently diagnosed patients, increasing the likelihood of capturing initial pathological changes. 2) Despite inconsistencies in the literature, we also hypothesised that, given the nature of the signal to be analysed— primarily ERPs derived from EEG, spatially informed by fMRI—and the existing ERP findings in MS, we would observe a predominance of weakened functional connections. This pattern would reflect the early impact of MS on neuronal communication, particularly in the form of slowed and less efficient processing.

## 2. Materials and methods

This study was approved by the ethics committees of the Faculty of Medicine of the University of Coimbra (reference CE-047/2018) and of the Centro Hospitalar e Universitário de Coimbra (CHUC) (reference CHUC-048-19) and was carried in accordance with the Code of Ethics of the World Medical Association (Declaration of Helsinki) for experiments involving humans. All participants gave written informed consent to participate in the study after a full verbal and written explanation. Data was handled according to the data protection laws in force. This study is reported according to the STROBE reporting guidelines for cohort studies.^34^

A schematic of the data analysis pipeline is shown in Fig.1A.

**Figure 1:**
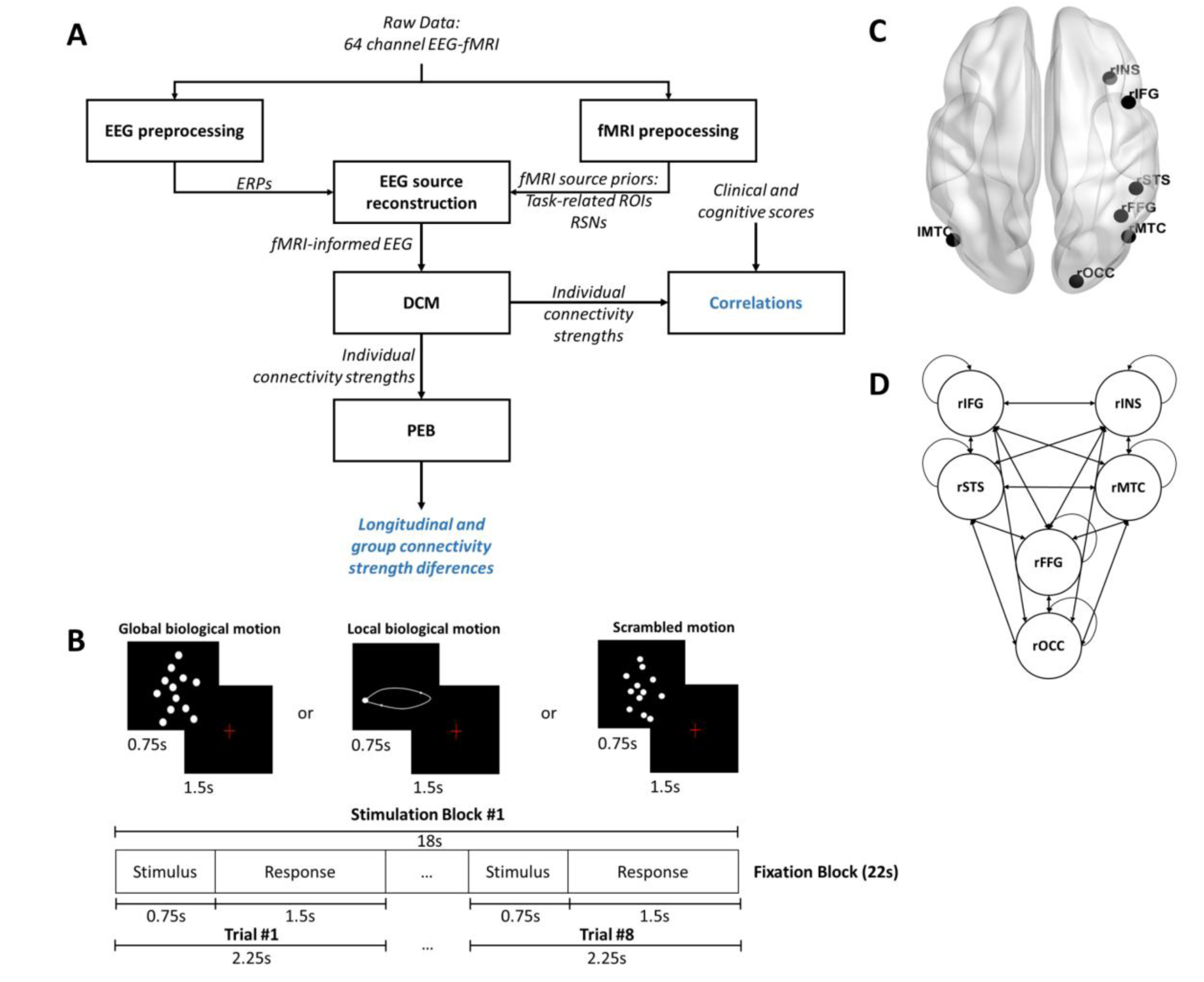
Overview of the analysis workflow, BM task protocol and BM network specification. (A) Schematic of the analysis pipeline, from simultaneous EEG-fMRI data collection, EEG and fMRI preprocessing, EEG source reconstruction, DCM specification, PEB analysis and correlations between connectivity measures and clinical and neuropsychological data. Boxes represent methods, italic represent input/output data of each method, blue represents the final outputs of the pipeline. (B) Experimental protocol for BM task. (C) Brain regions involved in the network activated by BM task and used for EC analysis. (D) Scheme of the network specification indicating connections between brain regions. DCM = dynamic causal modelling; ERP = event-related potential; fMRI = functional MRI; lMTC = left middle temporal cortex; PEB = parametric empirical bayes; rFFG = right fusiform gyrus; rIFG = right inferior frontal gyrus; rINS = right insula; rMTC = right middle temporal cortex, rOCC = right occipital cortex; ROI = region of interest; rSTS = right superior temporal sulcus.

### 2.1 Participants

Patients were recruited and assessed at the local hospital’s Neurology Department. Inclusion criteria were: RRMS diagnosis (McDonald criteria), ^46^ age between 18–55 years, disease duration < 5 years and evidence of disease activity. This means that participants were either treatment-naïve or had shown a suboptimal response to one previous treatment. This criterion ensured that all participants entered the study in a comparable clinical state, allowing for the longitudinal assessment of disease progression and treatment response. Treatment must be initiated/switched after the first MRI acquisition. The decision on the treatment prescription for each patient was independent of the study participation and made by the treating neurologist. Exclusion criteria included history of brain injury, neurological/psychiatric disorders, alcohol/drug abuse, recent antidepressant treatment (within 2 months), recent relapse or steroid use (within 4 weeks), and contraindications for MRI scanning. Age- and sex-matched HC were also recruited from the general population.

#### 2.1.1 Clinical, cognitive and visual evaluation

Patients underwent clinical and neuropsychological assessments at baseline and follow-up, conducted by a neurologist (SB) and a neuropsychologist (IS), respectively. Clinical assessment included the expanded disability status scale (EDSS),^47^ to assess neurological disability and motor function and the modified fatigue impact scale (MFIS),^48^ to measure fatigue and its impact in daily life. The neuropsychological assessment was done using the brief international cognitive assessment for MS (BICAMS), ^49^ and the reading the mind in the eyes (RME) test. ^50^ BICAMS comprises three tests: brief visuospatial memory test (BVMT) and california verbal learning test (CVLT),^49^ which evaluate verbal and visual memory deficits respectively, and symbol digit modalities test (SDMT),^51^ to evaluate information processing speed. RME is a social cognition test used to assess the ability to interpret emotions and mental states based on visual cues.^50^

Visual assessments were conducted by qualified clinicians at the Neurology Department in collaboration with expert ophthalmologists. Visual function was assessed in both groups at baseline and follow-up using standard ophthalmological examination. This included measurements of visual acuity (VA) and intraocular pressure (IOP) for the right eye (*Oculus Dexter*, OD) and left eye (*Oculus Sinister*, OS). VA was recorded using the decimal scale, where 1.0 represents normal VA (equivalent to 20/20 in Snellen notation) ^52,53^ with values <1.0 indicating reduced acuity and values >1.0 reflecting above-average visual performance. IOP was measured in mmHg, with normal physiological values typically ranging between 10 and 21 mmHg.^54^

### 2.2 Study design and experimental Protocol

The experimental protocol used in this study consists of two runs of a decision-making visual task of BM perception,^33,55^, which includes three conditions: global BM, local BM and scrambled motion ^23,56^. BM stimuli were built based on human motion capture data collected at 60 Hz, comprising 12 point-lights placed at the main joints of a male walker. Each BM run consisted of 12 blocks of 40 seconds (each subdivided in 18 seconds stimulation blocks and in 22 seconds fixation blocks): 4 or 5 blocks (depending on the starting block) of the point-light walker facing rightwards or leftwards (global BM), 4 or 5 blocks showing only the point-light located at the right ankle and moving rightwards of leftwards (local BM), and 3 blocks of point lights randomly positioned across the y axis, while maintaining their true trajectory across the x axis (scrambled motion). A total of 9 global, 9 local and 6 scrambled blocks were presented during the two BM perception runs. Each 18 seconds stimulation block is composed of 8 trials of 2.25 seconds subdivided in 0.75 seconds of global BM or local BM or scrambled motion stimulus followed by 1.5 seconds of a fixation cross. After each stimulus presentation, during fixation cross, the participants reported the direction of motion of the dots (left or right) by pressing one of two buttons of a response box. (Fig.1B). ^23,48^ Each BM run lasted approximately 8.37 minutes.

#### 2.2.1 Task Behaviour

For each trial of the BM task, response time (RT) and the accuracy measures, i.e., number of correct, incorrect, invalid, and unanswered trials were recorded for each participant. A correct trial was defined as one in which the participant accurately identified the direction of the stimulus motion (left or right) by pressing the corresponding button. An incorrect trial occurred when the participant pressed the opposite button relative to the stimulus direction. Invalid trials included instances where more than one button was pressed simultaneously or when an unrecognised trigger (i.e., neither left nor right) was registered. Unanswered trials referred to trials in which no response was provided.

### 2.3 EEG-fMRI data acquisition

Participants underwent imaging sessions at baseline (shortly after diagnosis) and follow-up (∼9.6 months later), using a 3T Siemens MAGNETOM Prisma Fit scanner (Siemens, Erlangen) and a 64-channel RF coil at the Portuguese Brain Imaging Network (Coimbra, Portugal). A reversed phase encoding direction (posterior-anterior) short acquisition was performed for image distortion correction. A 3D high-resolution anatomical image was also collected for subsequent functional imaging registration (Table 1). The EEG signal was recorded using a 64-channel NeuroScan SynAmps2 system with MaglinkTM software, and a cap of 64 non-magnetic Ag/AgCl electrodes arranged per the 10/10 system. A reference electrode was placed near Cz, and two electrodes were positioned on the back for ECG recording. Impedances were kept below 25 kΩ. EEG, ECG, and fMRI were acquired continuously, synchronized via Syncbox (NordicNeuroLab, USA), at a 10 kHz sampling rate and aligned with the MRI clock.

**Table 1.**
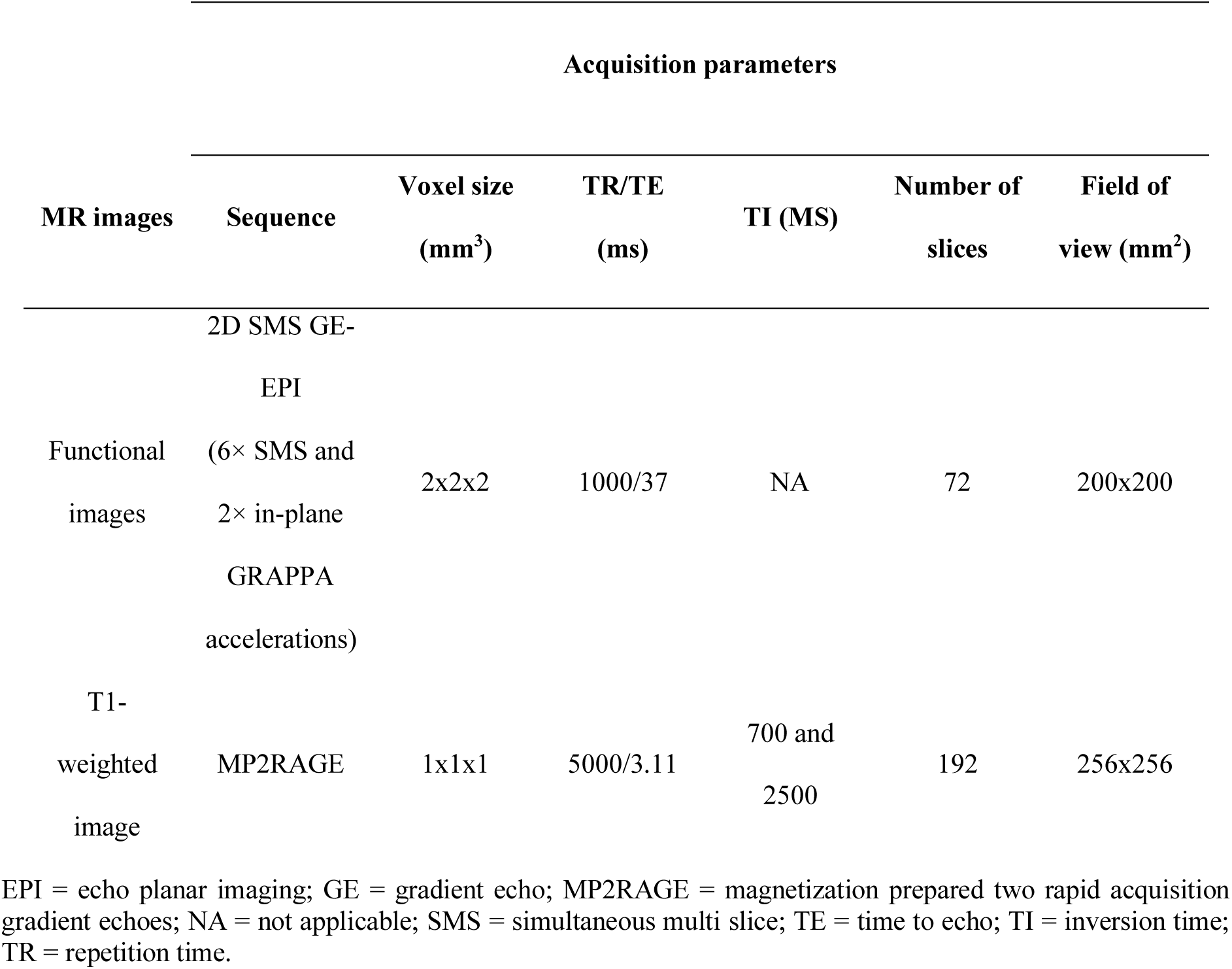
Acquisition parameters of the functional and structural MR images.

### 2.4 EEG preprocessing

EEG data were corrected for gradient artifacts using average artifact subtraction (AAS),^57^ implemented with FMRIB tools,^58^ —a plug-in of the EEGLAB toolbox,^59^, —in MATLAB (v.R2019b). Pulse artifacts were removed using the method presented by Abreu *et al.*^60^,: the EEG signal is first decomposed using independent component analysis (ICA), followed by AAS to the independent components (IC) associated with the artifact. The corrected signal is then reconstructed by combining the artifact-corrected ICs with the original non-artifact-related ICs. After, EEG preprocessing followed Cruz *et al*.^61^ pipeline: referencing to a robust estimate of the mean of all channels, bad channels and 1-second epochs (matching fMRI repetition time) removal, and an additional ICA for removing additional sources of EEG artifacts using the ICLabel algorithm, ^62^, implemented as a plug-in of the EEGLAB toolbox. EEG was then down-sampled to 500 Hz, band-pass filtered (0–45 Hz), segmented into stimulus-locked epochs (−200 to 800 ms), baseline-corrected, and averaged by condition to compute event-related potentials (ERP). Final data consisted of ERPs across 64 electrodes for each condition and participant.

### 2.5 fMRI data preprocessing

fMRI data were preprocessed using the SPM12 software (https://www.fil.ion.ucl.ac.uk/spm/software/spm12/)., with CAT12,^63^ PhysIO,^64^ and ArtRepair toolboxes,^65^ in MATLAB (v.R2019b), and the FMRIB Software Library (FSL)^58^ The preprocessing pipeline, described in Soares *et al*.^23^ included: slice timing, realignment, geometric distortion (FSL’s tool TOPUP^66^) and bias correction, image registration, segmentation of the T1-weighted sMRI to extract WM and ventricular CSF masks, nuisance regression (e.g., physiological signals, motion), motion spikes interpolation (ArtRepair^65^) spatial smoothing, temporal filtering, and normalization to Montreal Neurological Institute (MNI) standard space for consistent spatial alignment across subjects.^67^

### 2.6 fMRI priors for EEG source reconstruction

From the preprocessed fMRI data, resting-state networks (RSN), and task-related activity maps were extracted. These were the fMRI-derived priors used to constrain the EEG space through EEG source as detailed below

#### 2.6.1 Identification of resting-state networks

The preprocessed fMRI data were submitted to a group-level probabilistic spatial ICA decomposition using the FSL tool MELODIC^68^,whereby the data of each run for all participants is temporally concatenated prior to the ICA step, as recommended in the MELODIC’s guide for the identification of RSNs (https://fsl.fmrib.ox.ac.uk/fsl/fslwiki/MELODIC). The optimal number of ICs was automatically estimated based on the eigenspectrum of its covariance matrix. The spatial maps of the ICs were compared with those of the 10 RSN templates described in Smith *et al*.,^69^ using the dice coefficient to quantify spatial overlap.^70^ For each template, the IC map yielding the highest Dice coefficient was determined as the corresponding RSN. In the cases of non-mutually exclusive assignments, the optimal assignment was determined by randomizing the order of the RSN templates (a maximum of 10000 possible combinations were considered, for computational purposes), and then sequentially, and mutually exclusively, assigning them to the IC maps based on their Dice coefficient. The assignment with the highest average Dice coefficient across all RSN templates was then deemed optimal yielding the final set of RSNs: three visual networks, the default mode network (DMN), a cerebellum network, a motor network, an auditory network, the salience network, a right language network and a left language network .Further details can be found in Abreu *et al.*^56^

#### 2.6.2 Task-related activity mapping

BM task-related regions (Fig.1C) were mapped using a general linear model (GLM) with three regressors: global BM, local BM, and scrambled motion. To minimize bias toward specific BM in building fMRI priors for EEG source reconstruction, we identified visual motion-related regions using the contrast [global BM + local BM + scrambled motion – baseline]. Significance was defined as cluster-level *p* < 0.05, corrected for multiple comparisons using family-wise error (FWE) based on random field theory (RFT). One GLM with functional images from the two BM runs was estimated for each participant. Group-level analyses were then performed separately for each group and time point (MS at baseline and follow-up; the same for HC).

### 2.7 EEG source reconstruction

EEG source reconstruction was performed in SPM12 software (https://www.fil.ion.ucl.ac.uk/spm/software/spm12/), to confirm that a priori selected sources for the connectivity model are indeed active during task performance. Source mapping began with a forward model using a realistic head model. sMRIs were segmented into brain, scalp, and skull, then co-registered to Montreal Neurological Institute space. Electrodes (10/10 system) were aligned to the scalp and manually adjusted to match the distortions observed on sMRI images. A realistically shaped volume conduction model was estimated using a boundary element model (BEM) with three layers (scalp, inner skull and outer skull). 8196 source dipoles were placed at the vertices of a MNI-derived cortical mesh, and the leadfield matrix was estimated by mapping each possible dipole configuration onto a scalp potential distribution. ERP data were converted to SPM12 format, downsampled to 90 Hz and fed to multiple sparse priors (MSP) source reconstruction algorithm allowing the integration of fMRI priors (RSNs and task activation maps),^71,72^ as detailed described in Abreu *et al*.^56^ Group source reconstruction was used to improve and constrain source locations across subjects, varying only in activation strength, as recommended.^73^ Source activity with a time window 0–750 ms, frequency window of interest 0–48 Hz and by employing a Hanning taper was computed per condition, group, and time point. Inversion results were smoothed (six iterations) and converted to 3D NIfTI in MNI space.

### 2.8 Dynamic Causal Modelling

DCM for evoked responses was implemented using SPM12 software ((https://www.fil.ion.ucl.ac.uk/spm/software/spm12/),to model the neuronal dynamics underlying BM through a convolutional neural mass model based on the three-cell population.^74,75^

#### 2.8.1 Network definition

As our focus of interest was the task-dependent modulation of EC (inference on model parameters), rather than comparison of different models (inference on model architecture), we used the already established BM processing network model from Sokolov *et al*.^33^ These regions were validated through our reconstructed source activation maps, ensuring consistency with the original framework. In line with this and based on the functional organization of brain networks and the hierarchy levels of brain regions,^76–78^ we placed the right occipital visual cortex (rOCC) at the bottom (lower-level function) and right inferior frontal gyrus (rIFG) and right insula (rINS) at the top (higher-level function) (Fig.1D). In DCM, EC between and within sources is defined using three core matrices: A represents the intrinsic (baseline) connectivity between regions which subdivides into A{1}—forward (bottom-up), A{2}—backward (top-down), and A{3}—lateral connections; B captures task-dependent modulations of these baseline connections; here, this was defined by contrasting responses to global BM and scrambled motion, based on Sokolov *et al*.^33^; C indicates the entry point of experimental (external) inputs—in this case, directed to the rOCC, reflecting early visual processing input.

#### 2.8.2 Model specification

ERPs (time windows: 0-750 ms) were fitted to the fully connected model specified above and activity of cortical sources was modeled using the equivalent current dipole (ECD) method.^74^ Each participant model was inverted according to a variational Bayesian scheme to examine the likelihood of parameters^79^ This approximates the posterior probability (Pp) of model parameters 𝑝(𝜃|*y, m*), i.e., the probability of the model parameters given the data and the model. The ERPs responses are represented by *y*, and *m* represents the model, i.e., which regions in the brain are connected and how these connections are modulated by the tasks; 𝜃 represents the model parameters. Each model yielded 47 parameters, which were statistically analysed for disease-related effects using PEB.

#### 2.8.3 Statistics over DCM parameters – Parametric Empirical Bayes

Statistical inference on the DCM parameters was done using PEB. It comprises a Bayesian GLM at the second level which collects all the subjects’ neural parameters, ordered by subject and then by parameter and describes them as being a linear combination of regressors in a design matrix (one column for each experimental effect, for instance the effect of group affiliation on one connection) and by group-level parameters.^80^ We focused the statistical inference on EC parameters modulated by experimental effects (matrix B) – global BM versus scrambled motion. The rationale for the following comparisons is grounded in a translational perspective. From a clinical standpoint, it is important to address the following questions: (1) How is connectivity altered in MS compared to HC shortly after diagnosis? (2) How is connectivity altered in MS compared to HC 10 months after diagnosis? (3) Are there longitudinal connectivity changes within the patient group? Finally, considering that connectivity changes also occur over time in healthy individuals due to natural and adaptive plasticity, (4) do these differ from connectivity changes observed in MS, potentially reflecting disease-specific rather than adaptive neuroplasticity?

To study group differences at baseline (MS vs. HC) and to assess connectivity 10 months post-diagnosis and treatment (MS at follow-up vs. HC at baseline), we used a Bayesian general linear model (GLM) with two regressors: the first captured the group mean (a column of ones), and the second coded group affiliation (1 for MS, -1 for HC). The model was applied to baseline data from both groups for the first comparison, and to follow-up data from MS and baseline data from HC for the second.

To examine longitudinal changes within the patient group (follow-up vs. baseline), we used the same GLM structure, with the time point coded as the second regressor (1 for follow-up, - 1 for baseline), applied only to patient data.

Finally, to test whether EC changes over time MS differed from those observed in HC, we used a three-level hierarchical model (PEB-of-PEBs).^80,81^ A second-level model was created for each group separately, with regressors for group mean and time point. These second-level parameters were entered into a third-level model with a 2×2 design matrix: one regressor for the overall mean and another for group difference (−1 for HC, 1 for MS). This model estimated parameters for overall mean connectivity, main effects of time point and group, and their interaction. All regressors were mean-centered.

For all PEB models, we used Bayesian model reduction followed by Bayesian model averaging. This essentially re-estimates model parameters at the level of individual DCMs by conducting a search over all possible parameter combinations that encode the design matrix. We reported the expected probability (Ep) of parameters to describe which connections in our models were strengthened/weakened as a result of disease (Supplementary Table 1 and 2) and over time (Supplementary Table 3) with strong evidence by Pp > 0.95.

### 2.9 Statistical analyses

#### 2.9.1 Clinical and cognitive evaluation

Clinical and cognitive measures in patients were summarised as median [min–max] or mean ± standard deviation (SD), depending on data distribution. Longitudinal comparisons (baseline vs follow-up) were performed using the Wilcoxon signed-rank test or the paired-samples *t*-test for non-normal and normal distributions, respectively.

#### 2.9.2 Visual assessment

Visual assessment was summarised as median [25th–75th percentiles] or mean ± SD for VA and IOP, separately for each eye and group, depending on data distribution. Group comparisons were performed using the Mann–Whitney U test or independent-samples *t*-test, depending on whether the data followed a non-normal or normal distribution, respectively. All analyses were conducted separately for the baseline and follow-up sessions.

#### 2.9.3 Task Behaviour

For each participant, the mean ± SD of RT were computed across all BM trials. Group-level RT was then summarised as mean ± SD. Accuracy measures were summarised as the total number of correct, incorrect, invalid, and unanswered trials for each participant. For each response category, the median and 25th–75th percentiles were calculated within each group. To assess group differences, independent samples *t*-tests were used for mean RTs, while the Mann–Whitney U test was applied to the response category counts. This was performed for baseline and follow-up.

#### 2.9.4 EEG sensor space analysis

EEG sensor-level analysis was conducted across all time points and channels using the LIMO EEG toolbox,^82^ part of the EEGLAB,^59^ integrated in MATLAB (v.R2019b). On trial-averaged ERP amplitudes, one-sample t-tests were used to identify task-related effects, and repeated measures ANOVAs to investigate potential group and longitudinal differences, specifically, to assess effects of *condition*, *group* and *time point.* Results are reported corrected for multiple testing using spatial-temporal clustering with a cluster forming threshold of *p < 0.05* (Supplementary Material).

#### 2.9.5 fMRI-informed EEG source maps

To map the sources involved in BM task, 3D images resulting from EEG source reconstruction for each condition and participant were entered into a three-way ANOVA *(group*: MS and HC; *condition*: global BM, local BM and scramble; *time point*: baseline and follow-up). To enhance robustness, *t*-statistics maps were generated with the contrast [global BM motion + local BM motion + scrambled motion - baseline], with significance at *p* < 0.05 FWE-corrected.

#### 2.9.6 Correlations between connectivity and clinical and cognitive scores

After estimating EC with DCM and PEB, we explored associations between EC strength of altered connections in the MS group and clinical and cognitive scores (EDSS, MFIS, BICAMS, RME) using Spearman correlations. This analysis was done for the two time points: baseline and follow-up. Outliers (1.5 times the IQR above the third quartile or below the first quartile) were excluded. To account for multiple comparisons, p-values were corrected with false discovery rate (FDR Benjamini-Hochberg).^83^

## 3. Results

### 3.1 Participants

We recruited 18 patients with RRMS and 18 HC. Participants performed a baseline and a follow-up session (mean time between visits 9.57 ± 0.49 months for patients and 9.70 ± 0.62 months for HC) with the same imaging acquisition parameters and protocols, except for five HC due to COVID-19 pandemic restrictions on the research center (Table 2). At baseline twelve patients were treatment naïve and the remaining that were on treatment presented a suboptimal response to a previous treatment, so they were considered as presenting disease activity. Right after the baseline session the 12 treatment naïve participants initiated treatment and the remaining switched the treatment. Only one patient changed the medication between baseline and follow-up, due to side effects.

**Table 2.**
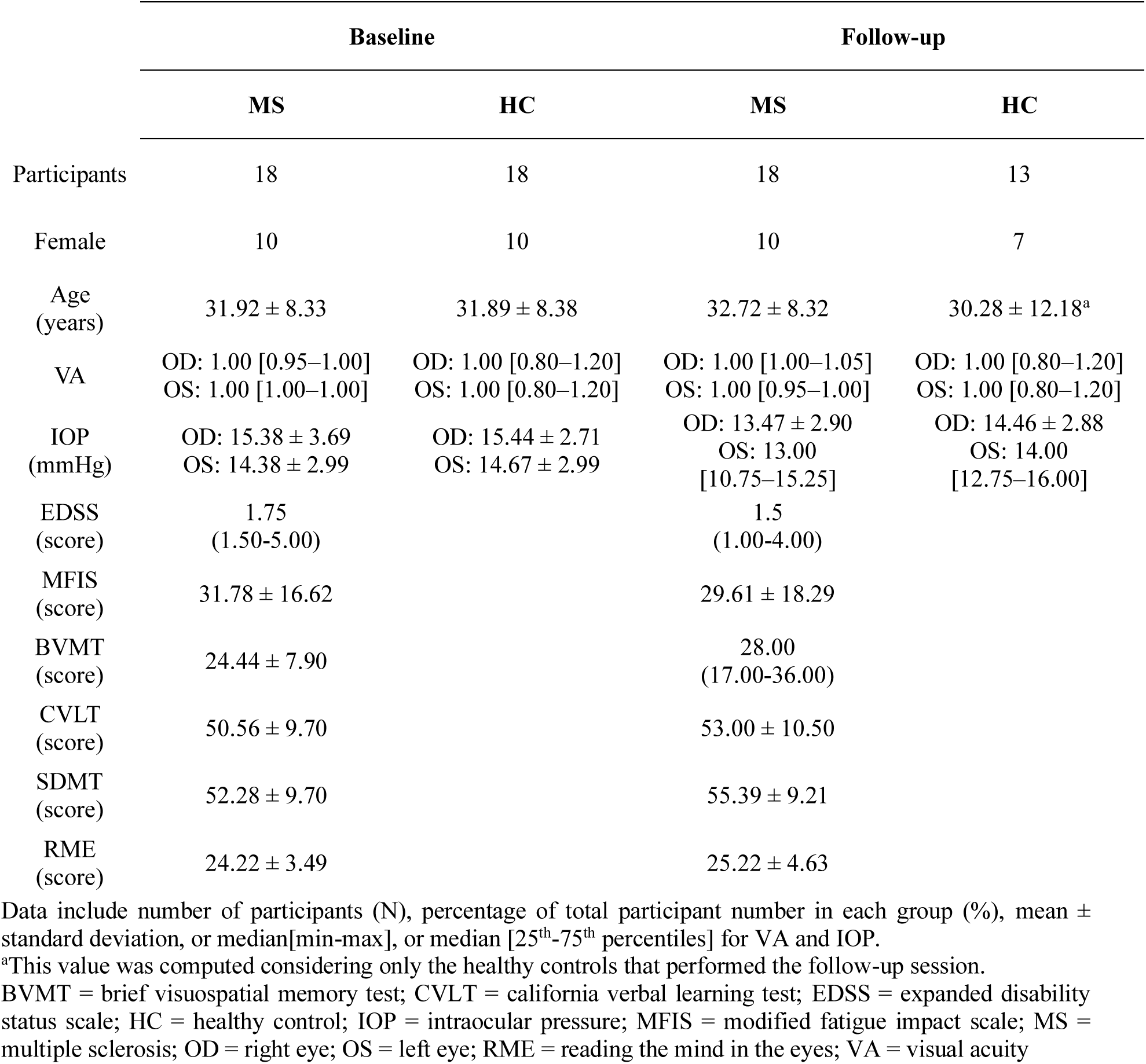
Descriptive statistics for demographic and clinical data.

#### 3.1.1 Clinical, cognitive evaluation

All clinical and cognitive scores showed improvement from baseline to follow-up, although these changes were not statistically significant — EDSS (*p* = 0.11), MFIS (*p* = 0.42), SDMT (*p* = 0.06), CVLT (*p* = 0.31), and RME (*p* = 0.16) — except for the BVMT (*p* = 0.03) (Table 2).

#### 3.1.2 Visual assessment

No significant group differences were observed in VA or IOP at baseline or follow-up. At baseline, comparisons between MS and HC yielded non-significant results for both eyes in VA (*p* = 0.42 for OD, *p* = 1.00 for OS) and IOP (*p* = 0.95 for OD, *p* = 0.78 for OS). At follow-up, VA remained comparable between groups (*p* = 0.84 for OD, *p* = 0.74 for OS), as did IOP (*p* = 0.36 for OD, *p* = 0.44 for OS) (Table 2).

Five patients had a history of optic neuritis, which was considered in the interpretation of visual outcomes. Only one patient showed slightly reduced values in both VA and IOP, falling below the normal range.

#### 3.1.3 Task Behaviour

At baseline, no significant differences in RT were found between MS (0.51 ± 0.14 s) and HC (0.44 ± 0.09 s; *p* = 0.11). Regarding accuracy, no incorrect or invalid responses were recorded. The number of unanswered trials did not differ significantly between groups (MS: 1.50 [0.00– 2.75]; HC: 2.00 [1.00–5.00]; *p* = 0.53). At follow-up, RT remained comparable between MS (0.47 ± 0.14 s) and HC (0.43 ± 0.09 s; *p* = 0.24). As at baseline, no incorrect or invalid responses were observed. The number of unanswered trials did not differ significantly between groups (MS: 2.00 [1.00–3.75]; HC: 4.00 [1.00–7.00]; *p* = 0.34).

### 3.2 EEG sensor space

No significant differences in all analysis of EEG sensor space were found (Supplementary Fig.3 and Fig.4) however, there’s a slight difference in amplitude—grand average time courses (mean across all participants)—for the global BM condition, between groups at the baseline and between time points in the MS group (Fig.2A).

**Figure 2:**
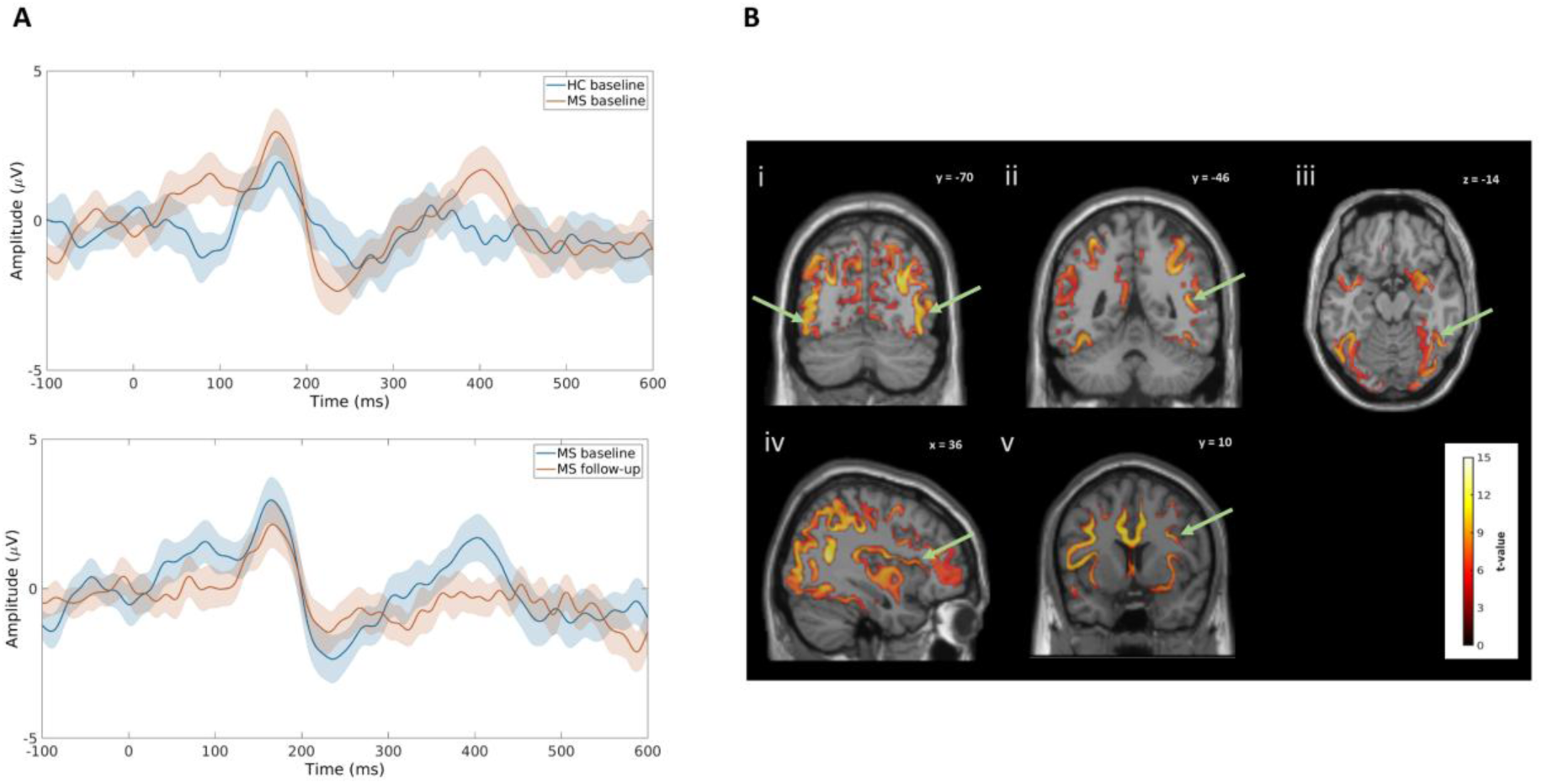
ERP group and longitudinal comparisons and fMRI-informed EEG source-level activation during BM perception. (A) Grand average ERP comparisons of global BM for a cluster of channels (P4, P6, P8, PO4, PO6, PO8, O2) between patients and HC at baseline (top) and between follow-up and baseline in patients group (bottom). (B) Brain source identification of brain regions involved in the BM task after EEG source reconstruction informed by fMRI priors. Colour bar indicates t-values scores. Green arrows represent the location of the regions previously identified by Sokolov et.al.41 i) bilateral middle temporal cortex (MTC; MNI: 46, -68, 0 and -48, -70, -2); ii) right superior temporal sulcus (rSTS; MNI: 50, -40, 10); iii) right fusiform gyrus (rFFG; MNI: 42, -56, -14); iv) right insula (rINS; MNI: 36, 24, 2); v) right inferior frontal gyrus (rIFG; MNI: 46, 10, 32). Activation of the right early visual occipital cortex (rOCC; MNI: 18, -94, 0) is also seen in iii and iv. BM = biological motion; HC = healthy controls; MS = multiple sclerosis.

**Figure 3:**
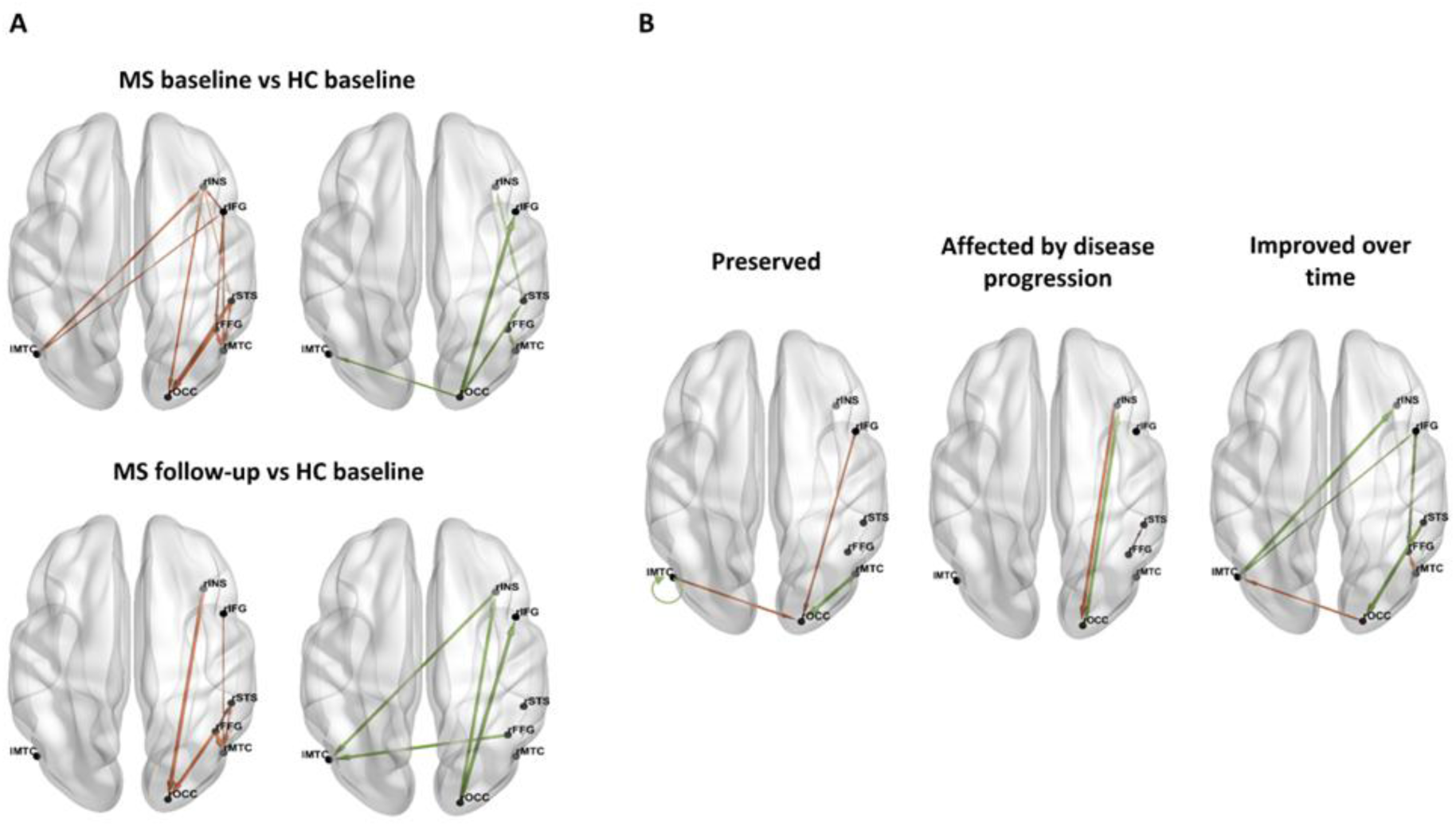
Group and longitudinal differences in EC. (A) EC differences between groups (MS and HC) at the two time points (baseline and follow-up). Differences between groups at baseline(top) and EC differences between MS at follow-up and HC at baseline (bottom) Red represents connections with decreased connectivity strength in MS relatively to HC. Green represents connections with increased connectivity strength in MS relatively to HC. (B) Longitudinal EC differences between time points (follow-up and baseline) in patients. Preserved connections (left), connections affected by disease progression (middle) and connections that improve over time (right). Red represents connections with decreased connectivity strength at follow-up relatively to baseline. Green represents connections with increased connectivity strength at follow-up relatively to baseline. Arrow width is proportional to connectivity strength difference HC = healthy controls; lMTC = left middle temporal cortex; MS = MS; rFFG = right fusiform gyrus; rIFG = right inferior frontal gyrus; rINS = right insula; rMTC = right middle temporal cortex; rOCC = right occipital cortex; rSTS = right superior temporal sulcus

**Figure 4:**
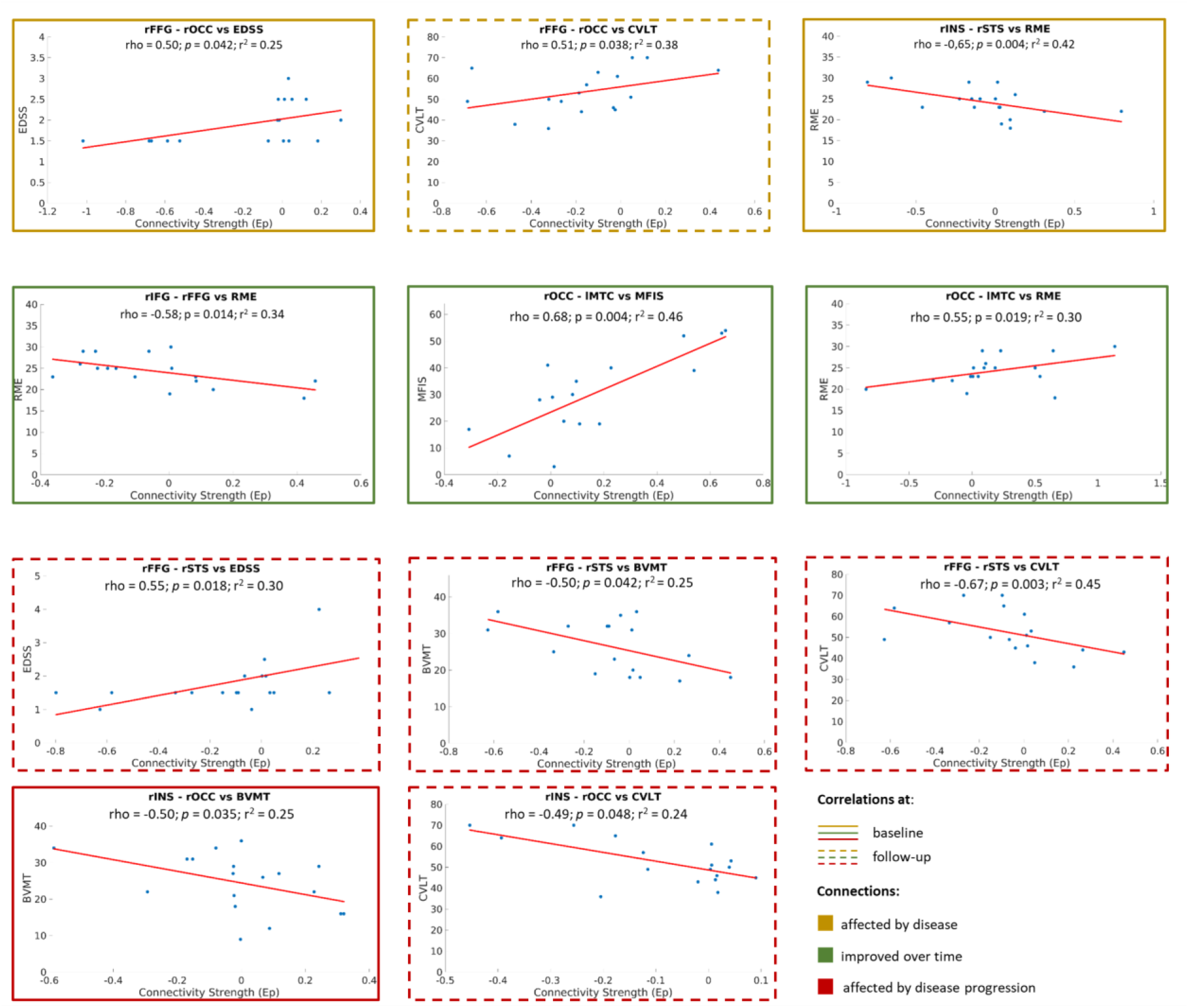
Scatter plots of correlations between connectivity strength of connections that are altered in patients and clinical and cognitive scores. Correlations with solid outlines appear at baseline, and correlations with dashed outlines appear at follow-up. Correlations with yellow outlines refer to connections that are affected by disease (altered at baseline and/or follow-up but do not change over time), correlations with red outlines refer to connections that are affected by disease progression, and correlations with green outlines refer to connections that improved over time. BVMT = brief visuospatial memory test; CVLT = california verbal learning test; EDSS = expanded disability status scale; Ep = expected probability; lMTC = left middle temporal cortex; rFFG = right fusiform gyrus; rIFG = right inferior frontal gyrus; rINS = right insula; RME = reading the mind in the eyes; rOCC = right occipital cortex; rSTS = right superior temporal sulcus.

### 3.3 Brain sources identification

We were able to identify the same brain regions as Sokolov and colleagues,^33^ in source activation maps. These are highlighted inFig.2B. The MNI coordinates of these regions were used in the specification of DCM models.

### 3.4 PEB

#### 3.4.1 EC alterations at baseline – MS vs. HC at baseline

16 connections demonstrated strong evidence (Pp > 0.95) of being different between groups at baseline. MS present 11 connections with decreased connectivity strength and five connections with increased connectivity strength comparatively to HC (Fig.3A; Supplementary Table 1).

#### 3.4.2 EC alterations at follow-up – MS at follow-up vs. HC at baseline

10 connections exhibited strong evidence (Pp > 0.95) of being different between MS at follow-up and HC at baseline. MS present six connections with decreased connectivity strength and four connections with increased connectivity strength comparatively to HC (Fig.3A; Supplementary Table 2).

#### 3.4.3 Longitudinal EC alterations in MS

13 connections demonstrated strong evidence (Pp > 0.95) of being different between time points in MS. At follow-up, seven connections show increased connectivity strength values, while six connections exhibit decreased connectivity strength compared to baseline (Table 3; Fig.3B; Supplementary Table 3).

**Table 3.** EC changes over time in MS.

| Connections | MS<sub>f</sub> vs MS<sub>b</sub> | MS vs HC |  |
| --- | --- | --- | --- |
|  |  | Baseline | Follow-up |
| rMTC – rOCC | MS<sub>f</sub> > MS<sub>b</sub> | No difference | No difference |
| lMTC – lMTC | MS<sub>f</sub> > MS<sub>b</sub> | No difference | No difference |
| lMTC – rOCC | MS<sub>f</sub> < MS<sub>b</sub> | No difference | No difference |
| rIFG – rOCC | MS<sub>f</sub> < MS<sub>b</sub> | No difference | No difference |
| rMTC – rFFG | MS<sub>f</sub> < MS<sub>b</sub> | MS > HC | No difference |
| lMTC – rIFG | MS<sub>f</sub> > MS<sub>b</sub> | MS < HC | No difference |
| lMTC – rINS | MS<sub>f</sub> > MS<sub>b</sub> | MS < HC | No difference |
| rSTS – rOCC | MS<sub>f</sub> > MS<sub>b</sub> | MS < HC | No difference |
| rIFG – rFFG | MS<sub>f</sub> > MS<sub>b</sub> | MS < HC | No difference |
| rOCC – lMTC | MS<sub>f</sub> < MS<sub>b</sub> | MS > HC | No difference |
| rOCC – rINS | MS<sub>f</sub> > MS<sub>b</sub> | No difference | MS > HC |
| rINS – rOCC | MS<sub>f</sub> < MS<sub>b</sub> | MS < HC | MS < HC |
| rFFG-rSTS<sup>a</sup> | MS<sub>f</sub> < MS<sub>b</sub> | No difference | MS < HC |

<sup>a</sup>Although this connection does not show significant change over time, it differs between groups at follow-up but not at baseline, suggesting an evolving pattern over time.
HC = healthy controls; lMTC = left middle temporal cortex; MS = multiple sclerosis; MS<sub>b</sub> = multiple sclerosis at baseline; MS<sub>f</sub> = multiple sclerosis at follow-up; rFFG = right fusiform gyrus; rIFG = right inferior frontal gyrus; rINS = right insula; rMTC = right middle temporal cortex; rOCC = right occipital cortex; rSTS = right superior temporal sulcus.

Considering how they differ between groups at each time point and to facilitate comprehension and interpretation we grouped these connections as: 1) preserved connections, if they show no differences between groups at any time point; 2) connections affected by disease progression, if they differ between groups at follow-up (unaffected at baseline or altered at baseline and follow-up; 3) connections improved over time if they differ between groups only at baseline. These connections approximate healthy values within the observed timeframe. While it remains uncertain whether this reflects a definitive improvement, we will refer to these connections as “improved” for the sake of simplicity (Table 3). In 2), two possible scenarios emerge: the connection was similar in both groups at baseline but changed over time or the connection was already impaired at baseline and remained as affected or even more at follow-up. While there are connections affected by disease, i.e., are altered at baseline and/or at follow-up compared to HC at baseline these are different from those considered as affected by progression of the disease because they change over time.

##### Connections that may be preserved

The connections rMTC-rOCC, lMTC-lMTC, lMTC-rOCC and rIFG-rOCC change over time but show no significant differences relative to HC at any time point. Specifically, rMTC-rOCC and lMTC-lMTC exhibit increased connectivity at follow-up, whereas lMTC-rOCC and rIFG-rOCC show decreased connectivity at follow-up, relative to baseline (Fig.3B).

##### Connections that may be affected by disease progression

The rOCC–rINS and rFFG–rSTS connections do not differ between groups at baseline but show alterations over time and differ between groups at follow-up. In MS, rOCC–rINS connectivity increases over time, becoming significantly higher than in HC at follow-up. In contrast, rFFG–rSTS connectivity decreases in MS (though not significantly), and at follow-up is significantly lower than in HC. While rFFG–rSTS does not show a significant time effect, the emerging group difference at follow-up suggests an evolving disease-related pattern. On the other hand, rINS-rOCC connectivity is significantly different from HC at baseline, presenting lower connectivity strength. Over time, connectivity decreases and at follow-up, and connectivity values are even lower in MS compared to HC. These are the two cases possible when considering connections affected by disease progression: a connection that was similar between both groups at baseline but changed over time (rOCC-rINS and rFFG-rSTS); or a connection was already impaired in MS at baseline and became more affected at follow-up, with disease continuing to impact it over time (rINS-rOCC) (Fig.3B).

##### Connections that may be improved

The connectivity strength of the lMTC – rIFG, lMTC – rINS, rSTS – rOCC, and rIFG – rFFG connections is significantly decreased in MS compared to HC at baseline. Over time, connectivity increases and at follow-up there are no significant differences compared to HC. On the other hand, rOCC-lMTC and rMTC-rFFG connectivity strength is significantly increased in MS compared to HC at baseline. Over time connectivity decreases, and at follow-up, there are no significant differences compared to HC (Fig.3B).

#### 3.4.4 Longitudinal EC trajectories (MS vs. HC)

The PEB results revealed four connections with strong evidence (Pp > 0.95) of changing differently over time between groups (Table 4).

**Table 4.**
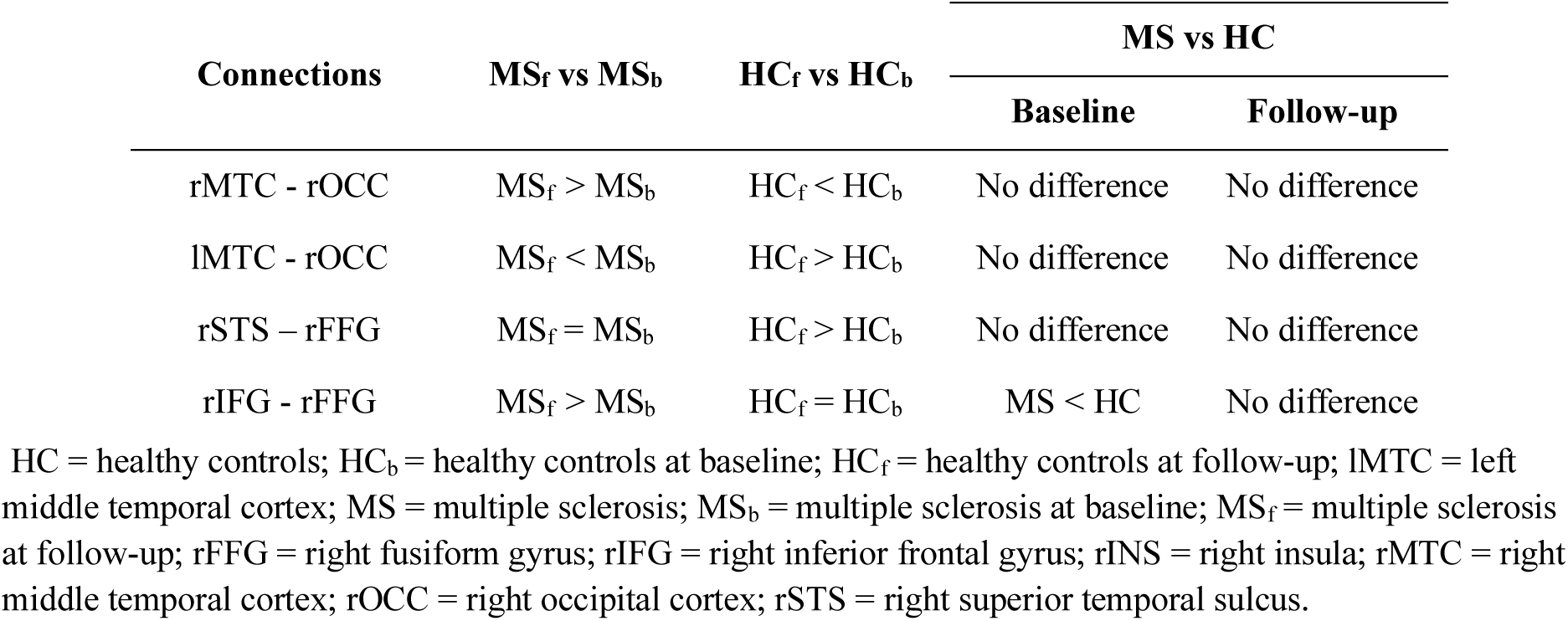
Connections that change over time in MS patients differently from HC.

The first two connections were categorized as preserved, at least in the time frame of this study, as they show no differences between groups at any time point, however they follow opposite directions regarding alterations over time compared to HC. The third connection, rSTS-rFFG, does not change significantly over time in MS, and does not differ from HC (comparison between MS and HC) at any time point. However, the tendency in HC is to increase over time. The fourth connection, rIFG-rFFG, is a connection that apparently improves over time as it differs from HC at baseline but over time connectivity in MS increases and at follow-up shows no differences compared to HC. In contrast, in HC, connectivity strength remains stable over time.

### 3.5 Correlations between connectivity and clinical and cognitive scores

Significant correlations between the strength of altered connections and clinical or cognitive scores are shown in Fig.4. No *p*-values survived multiple comparisons correction but to avoid overlooking potentially meaningful findings, we reported significant results at uncorrected *p* < 0.05.

## 4. Discussion

In this study, we investigated both cross-sectional and longitudinal changes in EC in MS during performance of BM task using simultaneous EEG-fMRI data. Additionally, we studied associations between connectivity strength and clinical measures of disability and cognition. The main findings of this work were the significant group differences and longitudinal changes in EC, along with robust associations between altered connections in MS and cognitive performance, namely social cognition, and physical disability. Specifically, some connectivity changes over time appeared to reflect treatment effects or adaptive plasticity, while others indicated ongoing disease progression. These results highlight the potential of task-based EC analysis using simultaneous EEG-fMRI to identify brain regions vulnerable to future decline, offering valuable potential biomarkers for predicting disease progression, symptom development, and therapeutic targets. These and other findings are further explored in the following structured sections.

### 4.1 Task Performance and visual function: ensuring comparable conditions across groups

The absence of statistically significant differences in visual function between groups, with both groups exhibiting values within the normal healthy range, indicates that participants were able to engage with the BM task without visual impairment and that task performance was not confounded by visual dysfunction. Similarly, no significant differences in task performance were observed between groups at any time point, indicating that both groups engaged with the task equally and effectively. These findings support the interpretation that the EC alterations reported in the subsequent sections are not confounded by differences in task performance or visual capacity but rather reflect disease-related functional changes.

### 4.2 EC alterations at baseline and associations with cognition and disability

Our results suggest that EC is already disrupted in early MS, with a predominance of weakened connections compared to HC, at baseline. This is in line with our second hypothesis, supporting the notion that weakened connectivity may reflect impaired functional integration. These altered connections—whether increased or decreased—impact both physical and cognitive function, as shown by positive correlations with EDSS (rFFG-rOCC) and MFIS (rOCC-lMTC), and negative correlations with RME (rINS-rSTS, rIFG-rFFG), and BVMT (rINS-rOCC). Although resting-state studies often found increased connectivity early on,^11,13,84^ task-based studies (using different task paradigms and metrics) associated cognitive decline to both increases and decreases in connectivity.^85–88^ Notably, the consistent emergence of weakened connections in our results supports our hypothesis and further underscores the power of task-based paradigms to uncover alterations that may remain undetected at rest, providing complementary and functionally specific insights into disease impact.

Still, adaptive compensatory mechanisms seem present at baseline, as shown by increased connectivity in rOCC-lMTC, which correlates positively with social cognition (RME scores). Yet, this may come at the cost of greater fatigue, given rOCC-lMTC positive correlation with MFIS scores. Similar patterns—like hyperactivation, increased FC, and ERP amplitudes, ^11,89–91^—have been reported in early RRMS and associated to preserved cognition but also more fatigue, especially when emotion-related regions are involved.^92–94^ Although rOCC and lMTC are not core emotion/social areas, here they belong to a broader emotion-related network. These findings suggest that fatigue and social cognition are associated through alterations in a common network, which reinforces the MS impact on both fatigue and social cognition.

It is important to consider that these alterations and their correlations with EDSS and cognitive scores may mainly reflect early-stage inflammation. Acute inflammation may drive early symptom onset—though its exact mechanisms remain unclear—as evidenced by symptom worsening during relapses, when inflammation peaks.^95,96^ Here, associations at baseline—like rINS-rSTS with RME and rFFG-rOCC with EDSS— and their subsequent disappearance at follow-up, even though they did not change significantly over time, supports that inflammation, rather than connectivity alterations, may underlie these early symptoms. This is further supported by evidence from the literature showing that treatment reduces inflammation to negligible levels at follow-up.^97^

### 4.3 EC alterations at follow-up and over time and associations with cognition and disability

Over time, there seems to be an attempt to restore connectivity to healthy values as suggested by the decreasing number of alterations at follow-up (comparison between groups) and by the number of improved connections over time— change over time and differ between groups at baseline only—versus the number of connections affected by disease progression— change over time and differ between groups at follow-up. This shift towards healthier values may reflect reduced inflammation that naturally occurs over time or due to treatment: a few studies have reported functional reorganization and symptom improvement following disease-modifying treatments. ^97,98^ This improvement is not reflected by positive correlations with cognition or negative correlations with physical disability at follow-up. Yet, although not significant, MS seemed to improve as they scored better clinical and cognitive scores at follow-up. Here, rOCC-lMTC and rIFG-rFFG stand out as indicators of potential positive treatment effects as they appear affected and correlate negatively with cognition at baseline, change over time and at follow-up show no significant differences between groups.

In fact, most of the correlations observed at follow-up involved connections affected by disease progression. These showed negative associations with cognitive performance (rFFG–rSTS with BVMT and CVLT and rINS–rOCC with CVLT), suggesting maladaptive compensation. Despite minimal patients’ disability, and a short interval between visits, these connections may already reflect early effects of neurodegeneration, making them potential markers of disease progression.

The only association observed at follow-up indicating an attempt to an adaptive compensatory mechanism is between rFFG-rOCC (altered at both time points but does not change over time) and CVLT, where higher connectivity correlated with better performance. Although these regions are primarily involved in visual and face processing,^33^ rather than verbal memory, these associations suggest they may be recruited to support cognitive functions when traditional networks are compromised as has been observed in previous studies.^45^

### 4.4 Distinct longitudinal connectivity trajectories in MS and HC

Connectivity changes over time in HC may reflect natural and adaptive neuroplasticity. The interaction analysis aims to determine whether the longitudinal changes observed in the patient group follow a similar trajectory. If so, such changes may reflect healthy plasticity mechanisms rather than being solely attributed to disease-specific pathological processes. Considering this, another indicator of connectivity improvement over time comes from this analysis, which identified four connections that changed over time in MS differently than in HC: rMTC–rOCC, lMTC–rOCC, rSTS–rFFG, and rIFG–rFFG. These connections were previously categorized as: preserved (they change over time but do not differ from HC at any time point); or improved over time. Although these connections are not currently considered as affected by disease or by disease progression, their evolution suggests they could still diverge from healthy patterns over time. This analysis helps differentiate, within this time frame, between connections that are recovering (possibly due to treatment), those that remain unaffected, and those that appear stable but may later deteriorate. However, these uncertainties highlight the importance of longer follow-up studies to monitor disease progression and gain clarity on the long-term impact.

### 4.5 Regions involved in the associations with cognition and disability

We used a BM task previously employed by Sokolov *et al.*^33^ to investigate connectivity in MS. This task targets highly myelinated brain regions,^34,35^ which are more susceptible to damage, making it sensitive to early FC changes. It also engages areas involved in multiple cognitive domains, especially social cognition, which is often impaired in MS.^10,37,38^

In fact, connectivity strength in rINS–rSTS, rIFG–rFFG, and rOCC–lMTC connections was associated with RME scores. Notably, rINS-rSTS and rIFG-rFFG connections involve regions strongly and specifically associated to social cognition: rINS is key for empathy and risk perception,^99^ rSTS for theory of mind,^100^ (directly assessed by the RME test), rIFG for understanding others’ emotions and intentions,^101^ and rFFG for face recognition.^102^ These findings provide direct support for our primary hypothesis, suggesting that this paradigm approach can detect functionally meaningful alterations associated with social cognitive performance in MS. Interestingly, the only positive correlation with RME involved rOCC– lMTC, a connection with increased connectivity strength and primarily associated to visual motion perception rather than social cognition. In contrast, the other associations involve connections whose strength is decreased relative to HC. This pattern suggests a compensatory mechanism, where non-social regions may support cognitive function when core social networks are compromised. This is supported by previous studies which have reported increased FC in regions not typically associated with the target cognitive domain.^45^ These associations further emphasize the potential contribution of these connections to early social cognition-related symptoms, with rINS-rSTS presenting the strongest correlation (*rho* = -0.65). Moreover, they stress the importance of including RME in neuropsychological assessments to better detect subtle deficits and provide earlier interventions.

Overall, the BM task effectively engaged brain regions associated with cognitive domains assessed by BICAMS as evidenced by associations between EC and BVMT or CVLT scores in rFFG–rOCC, rFFG–rSTS, and rINS–rOCC connections. Additionally, correlations with EDSS—despite its focus on physical disability,^103^ —were observed in rFFG–rOCC and rFFG–rSTS. These findings highlight the ability of this task paradigm, and targeted task paradigms in general, and these neuroimaging techniques to identify early symptom-specific alterations.

No significant associations between SDMT performance and the connectivity strength of altered connections were found. It is possible that compensatory mechanisms are at play, temporarily preserving SDMT performance despite underlying neural disruptions. Alternatively, individual variability in disease progression and the specific regions affected may account for the absence of significant associations.

### 4.6 Task EEG-fMRI in detecting potential biomarkers

ERP analyses (Supplementary Fig.1 and Fig.2) showed consistent parieto-occipital activity between 130–200 ms post-stimulus, reflecting the P150 component of early visual processing.^104,105^ Source maps from fMRI-informed EEG confirmed the paradigm’s spatial specificity. Together, these findings underscore the sensitivity of this paradigm and support its use for investigating source-level connectivity changes.

Task-based paradigms may reveal distinct alterations, mitigated during rest, as previously suggested.^24,25^ In fact, we found that most of the significant correlations reflecting maladaptive mechanisms—greater disability and poorer cognition—involved connections with decreased connectivity which might reveal a pattern where the weaker the connectivity strength the greater is symptom severity and disease progression. This contrasts with resting-state studies, which often report early increased FC to compensate for structural damage and associated with preserved function. ^11^

However, as discussed earlier, increased connectivity in non-social information processing regions (rOCC–lMTC) may reflect a compensatory response— helping to counteract social cognition deficits—but at the cost of increased fatigue, indicating both adaptive and maladaptive mechanisms. Similar patterns of increased connectivity in task-unrelated regions, such as the sensorimotor network (typically associated to motor function), have been associated with cognitive deficits.^45^ These dynamics may go unnoticed in resting-state studies, underscoring the value of task-based neuroimaging in revealing such adaptations.

Moreover, the relationship between decreased connectivity and symptom severity highlights potential targets for cognitive rehabilitation. Strengthening these networks may help mitigate cognitive decline in MS. The approach and findings presented here may contribute to the first step towards rehabilitation: the identification of early targets.

This identification underscored both the network’s relevance and the sensitivity of EEG-fMRI integration. By estimating EC from high temporal precision EEG source signals constrained by fMRI-informed regions, we were able to detect subtle connectivity-level alterations not captured by ERPs activation metrics alone. These findings support our primary hypothesis, highlighting the added value of this multimodal approach in uncovering early and clinically meaningful neural changes in MS. Notably, certain altered connections stand out as specific indicators of disease progression— rFFG-rSTS and rINS-rOCC— and treatment efficacy— rOCC-lMTC. Specifically, rFFG-rSTS appears unaffected at baseline, showing no differences between groups, but becomes altered over time and correlates with cognition only at follow-up. On the other hand, rOCC-lMTC is a connection that was altered at the baseline and positively associated with MFIS and RME. However, over time, its connectivity returned to healthy values and followed a trajectory similar to that of HC, which might indicate a positive effect of the administered treatment. These alterations may not appear in all newly diagnosed patients or predict individual disease trajectories, nor do they fully explain all phenotypes given the diffuse nature of MS. To further support the clinical utility of these models large-scale Randomized Controlled Trials are needed to validate these findings, ideally using functional or EC biomarkers as outcome measures. However, the fact that we were able to observe these alterations over time shows the potential of functional EC metrics as biomarkers for early neurodegeneration, treatment monitoring, and identifying vulnerable regions for predicting progression and guiding interventions.

Moreover, the simultaneous acquisition of fMRI and EEG, allowed us to derive fMRI priors— such as activation maps—under the exact same conditions as the EEG data, ensuring that external factors do not confound the experimental setup. Future analyses should apply the same method separately to EEG and fMRI data. Comparing outcomes would reveal each modality’s unique value, especially EEG, a lower-cost tool with strong potential for clinical implementation. Importantly, EEG can be informed by prior fMRI data acquired in a single session and reused in follow-ups to monitor disease progression.

### 4.7 Limitations

This study is not without limitations, as is the modest sample size, which might have reduced statistical power. Recruitment was constrained by COVID-19 restrictions, and the chosen follow-up period may have missed gradual changes—though it captured transient effects relevant to disease progression. Despite this, the longitudinal design provided meaningful insights. To the best of our knowledge, only two prior studies applied DCM to MS, using either resting-state,^31^ or simple tasks,^32^ that may not fully capture the alterations associated with cognitive impairment. By using a task engaging higher-order regions—such as those involved in social cognition, this study offers novel and promising findings that can guide future research.

## Data Availability

All data produced in the present study are available upon reasonable request to the authors.

## CRediT authorship contribution statement

**Júlia F. Soares:** Conceptualization, Methodology, Formal analysis, Investigation, Writing - Original Draft, Writing – Review & Editing, Visualization. **Maria Caranova:** Investigation, Formal analysis, Writing - Review & Editing. **Irina Santos:** Investigation. **Sónia Batista:** Conceptualization, Investigation, Writing - Review & Editing. **Miguel Castelo-Branco:** Conceptualization, Resources, Writing - Review & Editing, Supervision, Funding acquisition. **João V. Duarte:** Conceptualization, Methodology, Investigation, Writing - Review & Editing, Supervision, Project administration, Funding acquisition

## Acknowledgements

We would like to thank the participants for taking the time to participate in this study. We are also very grateful to Sónia Afonso and Tânia Lopes for the help with MRI setup and scanning.

## Glossary

AAS: artifact template subtraction
BICAMS: brief international cognitive assessment for multiple sclerosis
BM: biological motion
BVMT: brief visuospatial memory test
CVLT: California verbal learning test
DCM: dynamic causal model
EC: effective connectivity
EDSS: expanded disability status scale
ERP: event-related potential
FC: functional connectivity
fMRI: functional MRI
GLM: general linear model
IC: independent component
ICA: independent component analysis
IOP: intraocular pressure
lMTC: left motor cortex
MFIS: modified fatigue impact scale
MNI: Montreal Neurological Institute
PEB: parametric empirical Bayes
Pp: posterior probability
rFFG: right fusiform gyrus
rIFG: right inferior frontal gyrus
rINS: right insula
RME: reading the mind in the eyes
rOCC: right occipital visual cortex
RRMS: relapsing-remitting multiple sclerosis
rSTS: right superior temporal sulcus
SDMT: symbol digit modalities test
sMRI: structural MRI.
VA: visual acuity

