## Supplementary Material for "EEG-fMRI reveals biological motion network disruptions as early markers of Multiple Sclerosis progression"

**EEG sensor space methods**

After computing averaged, time-locked EEG for each group, time point, and condition, instead of the traditional ERP approach—which typically focuses on selected time windows and electrodes—we performed mass-univariate linear modelling across all electrodes and time points. This approach provides a more comprehensive and statistically robust analysis of task-related EEG amplitude, allowing us to investigate group and longitudinal differences across the full 0–800 ms window and all channels using a hierarchical linear model approach, as implemented in the LIMO EEG toolbox (Pernet et al., 2011), an open-source toolbox integrated in EEGLAB (Delorme & Makeig, 2004).

We started by running a first level analysis (subjects’ level), by setting up a GLM with three categorical regressors corresponding to the three task conditions (global BM, local BM and scrambled motion). In these first level analyses, all trials for each time point and channel were modelled as the sum of a constant term and coefficients for the three experimental conditions expressed through the GLM:

$$Y = XB + \varepsilon$$

Where $Y \in R^{p\times n}$, is a matrix representing the observed EEG data with *p*  trials and *n* time points, $X \in R^{p \times m}$ is the design matrix, with *m* regressors coding for the three task conditions and a constant term (intercept), $B\in\mathbb{R}^{m\times n}$ is the matrix of estimated regression parameters (i.e. beta weights over time) and $\varepsilon\in R^{p \times n}$ is the residual error matrix. Parameter estimates were obtained using ordinary least squares (Pernet et al., 2011).

At the second-level (group-level) analysis, six statistical models were implemented. First, two one-sample *t*-tests were conducted on the trial-averaged EEG amplitudes to identify when and where EEG responses significantly differed from zero across all participants, separately at baseline and follow-up. Next, to assess group differences, two mixed ANOVAs were conducted at each time point (baseline and follow-up), with *group* (HC vs. MS) as the between-subjects factor and *condition* (global BM vs. scrambled motion) as the within-subject factor. Finally, assess changes over time, two longitudinal repeated-measures ANOVAs were performed within each group separately: one in the MS group and one in the HC group. In both cases, *time* (follow-up vs. baseline) and *condition* (global BM vs. scrambled motion) were entered as within-subject factors. Here, we only compared global BM and scrambled motion because these are the two “extreme” conditions necessary to isolate BM specific effects. Second-level (group-level) analyses were performed using robust statistics based on trimmed means, which reduce the influence of outliers and violations of normality (Pernet et al., 2011; Wilcox, 2014). For follow-up comparisons, MS participants at follow-up were compared to HC at baseline. This decision was driven by the limited number of HC with follow-up EEG data (n = 13) and the higher susceptibility of EEG to noise, inter-session variability, and lower signal-to-noise ratio compared to, for instance, fMRI data. Under these conditions, group-level analyses benefit from a more stable and balanced control reference.

Results are reported corrected for multiple testing using spatial-temporal clustering with a cluster forming threshold of *p* = 0.05. Effect sizes for each statistical analysis were calculated as maximum and median effect sizes within the significant spatiotemporal clusters. If the statistical comparison resulted in no significant cluster, then effect sizes were calculated as maximum and median effect sizes within the whole analysis window. Cohen’s d was used for one-sample t-test analyses, partial eta squared (ηp^2^) was used for group effects on repeated measures ANOVA, and Mahalanobis distance squared (D^2^) was used for within-subjects’ analyses and interactions on repeated measures ANOVA (Lakens, 2013).

### EEG sensor space results

#### Activations at baseline

One-sample t-test across all time points and EEG channels, combining multiple sclerosis and healthy controls groups as well as all conditions at baseline revealed that the signal amplitude was significantly different from zero in two spatiotemporal clusters: cluster 1 starting at 130ms and ending at 194ms, *p*-value = 0.02, effect size max *d* = 1.29, median *d* = 0.66 with average amplitude of 1.69 ± 0.62 μV; cluster 2 starting at 201ms and ending at 688ms, *p*-value = 1.0 x 10^-3^, effect size max *d* = 1.85, median *d* = 0.58 with average amplitude of 0.31 ± 1.29 μV. EEG scalp topographies reveal a significant cluster with a positive potential localized in the right parietal and occipital channels (cluster 1) along with a second cluster (cluster 2) displaying more widespread activation across the entire brain, encompassing all channels.

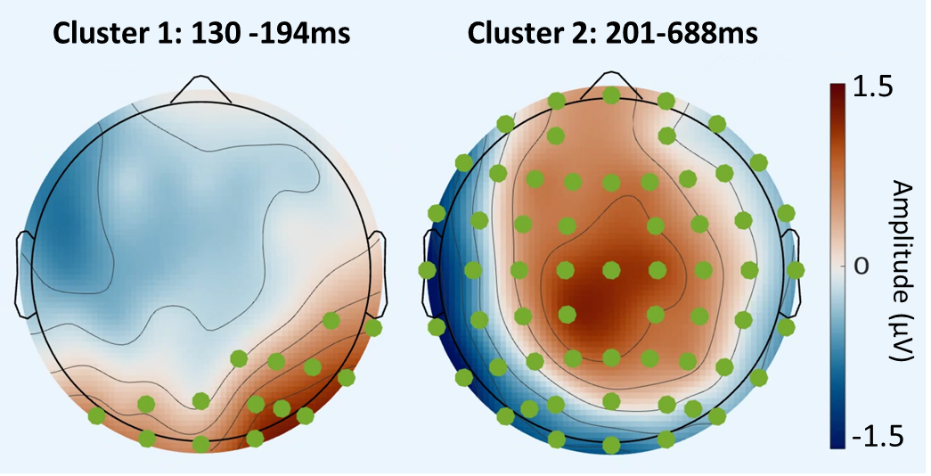

**Figure 1:** Scalp topographies of the grand average of EEG signal at baseline averaged within the time windows 130-194ms and 201-688ms. Green circles highlight the channels that belong to the significant spatiotemporal cluster where the EEG signal is significantly different from zero. The color bar represents amplitude values.

#### Activations at follow-up

One-sample t-test across all time points and EEG channels, combining patients and healthy controls groups as well as all conditions at follow-up revealed that the signal amplitude was significantly different from zero in four spatiotemporal clusters: cluster 1 starting at 132ms and ending at 186ms, *p*-value = 0.02, effect size max *d* = 1.22, median *d* = 0.65 with average amplitude of 0.91 ± 1.11μV; cluster 2 starting at 198ms and ending at 416ms, *p*-value = 1.0 x 10^-3^, effect size max *d* = 1.69, median *d* = 0.60 with average amplitude of 0.20 ± 1.43μV; cluster 3 starting at 420ms and ending at 550ms, *p*-value = 1.0 x 10^-3^, effect size max *d* = 2.18, median *d* = 0.63 with average amplitude of 0.25 ± 1.19μV; cluster 4 starting at 670ms and ending at 762s, *p*-value = 0.04, effect size max *d* = 0.49, median *d* = 0.68 with average amplitude of -0.92 ± 0.23 μV; EEG scalp topographies reveal a significant cluster with positive potentials localized around parietal and occipital channels and negative potentials around middle and frontal channels, clusters displaying more widespread activation encompassing the majority of channels (cluster 3 and 4) and a cluster with a negative localized potential around parietal and occipital channels.

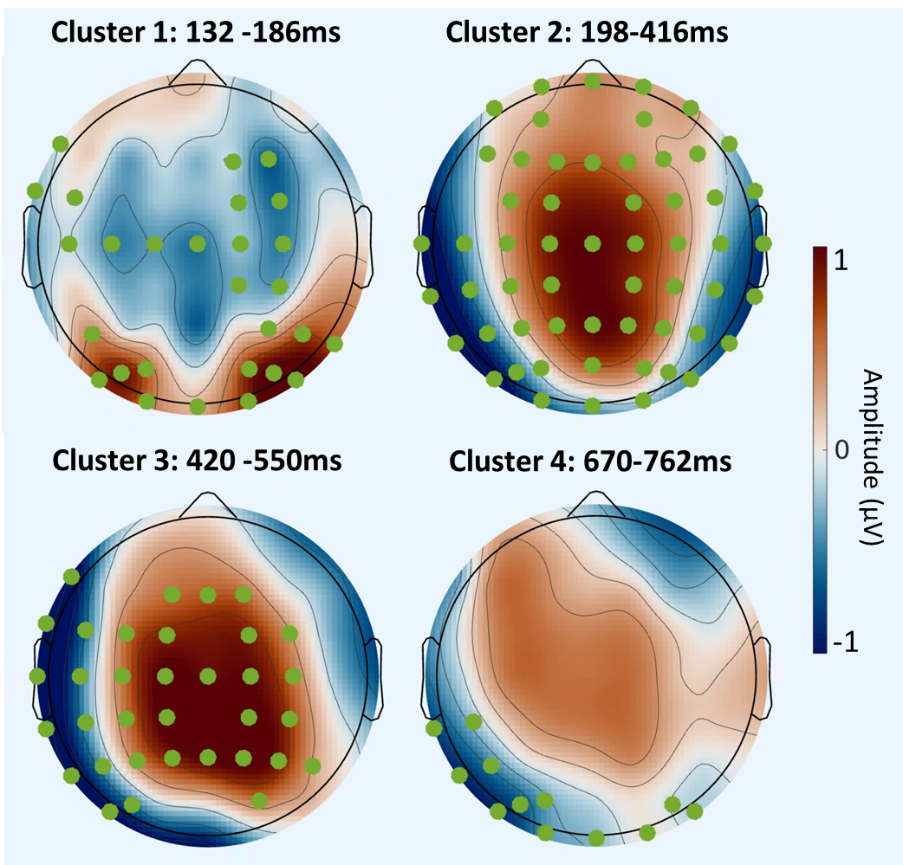

**Figure 2:** Scalp topographies of the grand average of EEG signal at follow-up averaged within the time windows 132-186ms, 198-416ms, 420-550ms and 670-762ms. Green circles highlight the channels that belong to the significant spatiotemporal cluster where the EEG signal is significantly different from zero. The color bar represents amplitude values.

These analyses show that the most localized activations appear to be at the parietal and occipital channels between 130 and 200ms.

#### Cross sectional analysis – differences between groups

Mixed ANOVA with group as between-subjects factor (MS vs. HC) and condition (global BM/scrambled motion) as within-subjects factor at baseline, revealed no significant group effect (effect size max ηp² = 0.30; median ηp² = 0.01), no significant effect of condition (effect size max D² = 3.7×10⁻³; median D² = 0.0001) and no significant effect of interaction between group and condition (effect size D² = 0.02; median D² = 4.0×10⁻⁴).

Mixed ANOVA with group as between-subjects factor (MS at follow-up vs. HC at baseline) and condition (global BM/scrambled motion) as within-subjects factor revealed no significant group effect (effect size max ηp² = 0.42; median ηp² = 0.01), no significant effect of condition (effect size max D² = 5.0×10⁻³; median D² = 1.0×10⁻⁴), and no significant effect of interaction between group and condition (effect size D² = 0.02; median D² = 4.0×10⁻⁴).

#### Longitudinal analysis – differences between time points

Repeated measures ANOVA with condition (global BM/scrambled motion) and time (patients at follow-up versus patients at baseline) as within-subjects factors revealed no significant effect (effect size max *D*^2^ = 0.34; median *D*^2^ = 0.01), no significant effect of condition (effect size max *D*^2^ = 5.0×10^−3^; median *D*^2^ = 1.0×10^−4^) and no significant effect of group x condition interaction (effect size *D*^2^ = 0.01; median *D*^2^ = 4.0×10^−4^).

Repeated measures ANOVA with condition (global BM/scrambled motion) and time (healthy controls at follow-up versus healthy controls at baseline) as within-subjects factors revealed no significant effect (effect size max *D*^2^ = 0.35; median *D*^2^ = 0.02), no significant effect of condition (effect size max *D*^2^ = 6.4×10^−3^; median *D*^2^ = 2.0×10^−4^) and no significant effect of group x condition interaction (effect size *D*^2^ = 0.03; median *D*^2^ = 6.0×10^−4^).

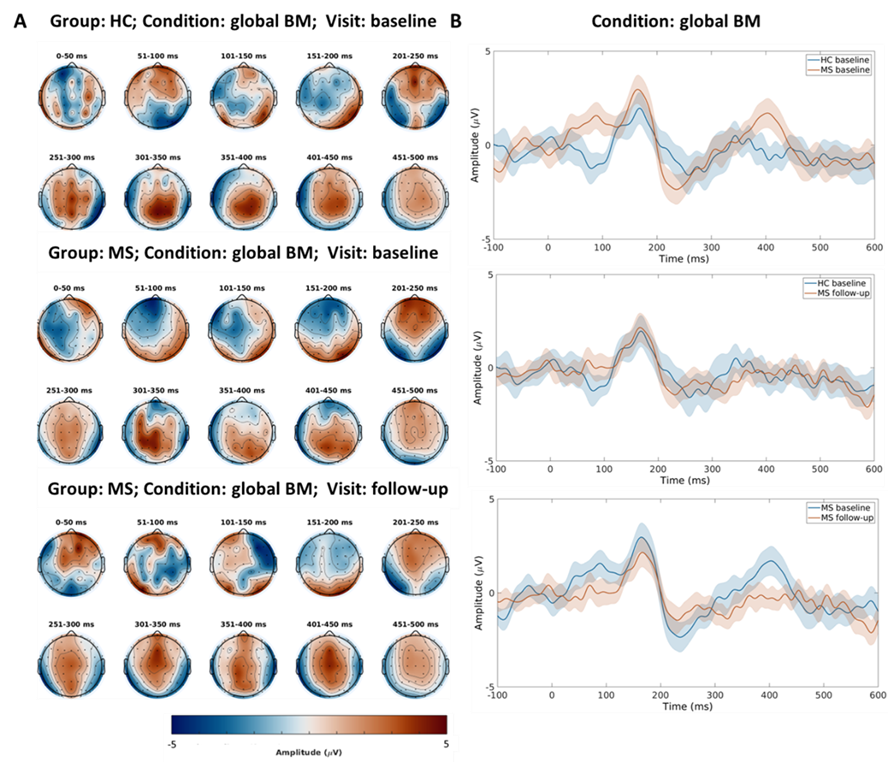

**Figure 3:** **EEG amplitude results.** **(A)** Scalp topographies from 0 to 600 ms for the global BM condition, depicting: the healthy controls group at baseline (first row), the patients group at baseline (second row), and the patients group at follow-up (third row). The color bar represents amplitude values. **(B)** BM grand average comparisons of a cluster of channels (P4, P6, P8, PO4, PO6, PO8, O2) between: healthy controls vs. patients at baseline (first row), healthy controls at baseline vs. patients at follow-up (second row), and patients at baseline vs. patients at follow-up (third row). Abbreviations: BM, biological motion; healthy controls, healthy controls; MS, multiple sclerosis.

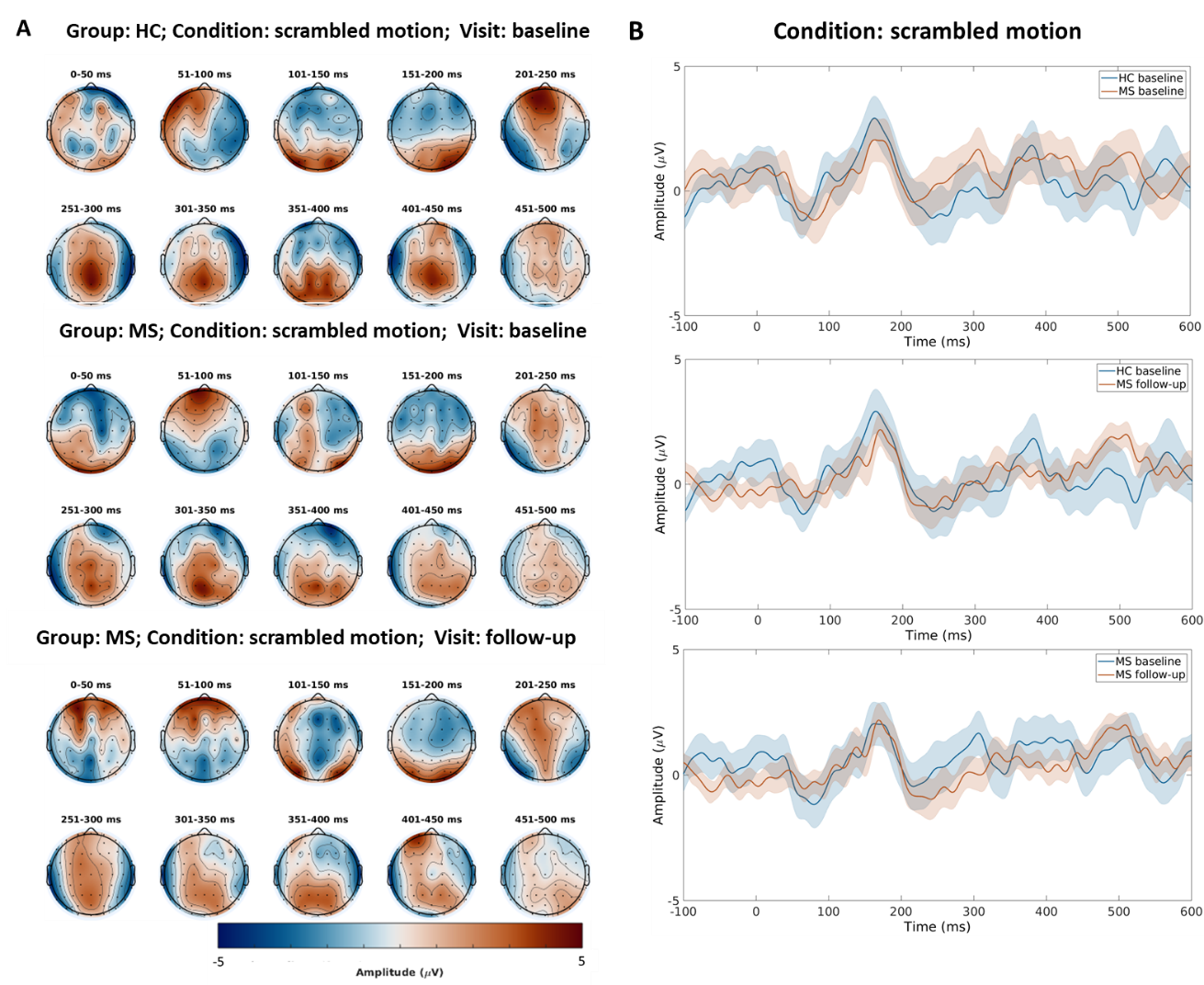

**Figure 4: EEG amplitude results. (A)** Scalp topographies from 0 to 500 ms for the scrambled condition, depicting: the healthy controls group at baseline (first row), the patients group at baseline (second row), and the patients group at follow-up (third row). The color bar represents amplitude values. **(B)** Scrambled motion grand average comparisons of a cluster of channels (P4, P6, P8, PO4, PO6, PO8, O2) between: healthy controls vs. patients at baseline (first row), healthy controls at baseline vs. patients at follow-up (second row), and patients at baseline vs. patients at follow-up (third row).

**PEB results**

##### EC alterations at baseline – MSvs. HC at baseline

**Table 1**: **PEB results of effective connectivity at the baseline.**

| **Connection** | **Ep (mean)** | **Pp (mean)** | **Ep (difference)** | **Pp (difference)** |
| --- | --- | --- | --- | --- |
| rFFG - rMTC | 0.201 | 1.00 | -0.241 | 1.00 |
| rFFG - rOCC | -0.001 | 0.00 | -0.333 | 1.00 |
| rMTC - rFFG | 0.276 | 1.00 | 0.155 | 1.00 |
| lMTC - rIFG | -0.138 | 1.00 | -0.147 | 1.00 |
| lMTC - rINS | -0.071 | 0.53 | -0.230 | 1.00 |
| rSTS - rMTC | -0.008 | 0.00 | -0.203 | 1.00 |
| rSTS - rINS | -0.007 | 0.00 | 0.190 | 1.00 |
| rSTS - rOCC | -0.371 | 1.00 | -0.339 | 1.00 |
| rIFG - rFFG | -0.066 | 0.54 | -0.172 | 1.00 |
| rIFG - rMTC | 0.009 | 0.00 | -0.153 | 1.00 |
| rIFG - rINS | 0.009 | 0.00 | -0.174 | 1.00 |
| rINS - rSTS | -0.001 | 0.00 | -0.139 | 1.00 |
| rINS - rOCC | 0.002 | 0.00 | -0.200 | 1.00 |
| rOCC - lMTC | 0.273 | 1.00 | 0.141 | 1.00 |
| rOCC - rSTS | -0.003 | 0.00 | 0.176 | 1.00 |
| rOCC - rIFG | -0.013 | 0.00 | 0.263 | 1.00 |

The values of the second and third columns are related to the mean EC across participants and the values of the fourth and fifth columns are related to the EC differences between patients and healthy controls. Ep = expected probability; Pp = posterior probability. rFFG = right fusiform gyrus; rIFG = right inferior frontal gyrus; rINS = right insula; rMTC = right middle temporal cortex; rOCC = right occipital cortex; rSTS = right superior temporal sulcus.

##### EC alterations at follow-up – MS at follow-up vs. HC at baseline

**Table 2: PEB results of effective connectivity at the follow-up.**

| **Connection** | **Ep (mean)** | **Pp (mean)** | **Ep (difference)** | **Pp (difference)** |
| --- | --- | --- | --- | --- |
| rFFG - rMTC | 0.182 | 1.00 | -0.290 | 1.00 |
| rFFG - lMTC | -0.007 | 0.00 | 0.157 | 1.00 |
| rFFG - rSTS | -0.013 | 0.00 | -0.193 | 1.00 |
| rFFG - rOCC | 0.002 | 0.00 | -0.281 | 1.00 |
| rSTS - rMTC | -0.007 | 0.00 | -0.194 | 1.00 |
| rIFG - rMTC | 0.126 | 1.00 | -0.111 | 1.00 |
| rINS - lMTC | 0.018 | 0.00 | 0.153 | 1.00 |
| rINS - rOCC | -0.136 | 1.00 | -0.313 | 1.00 |
| rOCC - rIFG | -0.134 | 1.00 | 0.191 | 1.00 |
| rOCC - rINS | 0.001 | 0.00 | 0.167 | 1.00 |

The values of the second and third columns are related to the mean EC across time points and the values of the fourth and fifth columns are related to the EC differences between follow-up and baseline in patients. Ep = expected probability; Pp = posterior probability. rFFG = right fusiform gyrus; rIFG = right inferior frontal gyrus; rINS = right insula; rMTC = right middle temporal cortex; rOCC = right occipital cortex; rSTS, = right superior temporal sulcus.

##### Longitudinal EC alterations in multiple sclerosis

**Table 3:** **PEB results of effective connectivity in patients.**

| **Connection** | **Ep (mean)** | **Pp (mean)** | **Ep (difference)** | **Pp (difference)** |
| --- | --- | --- | --- | --- |
| rMTC - rOCC | -0.208 | 1.00 | 0.285 | 1.00 |
| lMTC - lMTC | -0.012 | 0.00 | 0.170 | 1.00 |
| lMTC - rOCC | -0.004 | 0.00 | -0.167 | 1.00 |
| rIFG - rOCC | 0.009 | 0.00 | -0.169 | 1.00 |
| rMTC - rFFG | 0.289 | 1.00 | -0.176 | 1.00 |
| lMTC - rIFG | -0.171 | 1.00 | 0.141 | 1.00 |
| lMTC - rINS | -0.010 | 0.00 | 0.272 | 1.00 |
| rSTS - rOCC | -0.299 | 1.00 | 0.299 | 1.00 |
| rIFG - rFFG | 0.004 | 0.00 | 0.241 | 1.00 |
| rOCC - lMTC | 0.185 | 1.00 | -0.163 | 1.00 |
| rOCC - rINS | 0.010 | 0.00 | 0.146 | 1.00 |
| rINS - rOCC | -0.296 | 1.00 | -0.161 | 1.00 |
| rFFG-rSTS | -0,229 | 1,00 | -0,071 | 0,54^a^ |

The values of the second and third columns are related to the mean EC across time points and the values of the fourth and fifth columns are related to the EC differences between follow-up and baseline in patients. ^a^Although this connection does not show significant change over time, it differs between groups at follow-up but not at baseline, suggesting an evolving pattern over time

Ep = expected probability; Pp = posterior probability. rFFG = right fusiform gyrus; rIFG = right inferior frontal gyrus; rINS = right insula; rMTC = right middle temporal cortex; rOCC = right occipital cortex, rSTS = right superior temporal sulcus.
